# Longitudinal performance of the ENLIST ENL Severity Scale in individuals with severe erythema nodosum leprosum: responsiveness, trajectories and clinical features – a secondary analysis of the Methotrexate and Prednisolone study - MaPs in ENL

**DOI:** 10.64898/2026.05.26.26354110

**Authors:** Barbara de Barros, Abdulnaser Hamza, Alemtsehay Getachew, Medhi Denisa Alinda, Farha Sultana, Bishwanath Acharya, Vivek V. Pai, Anju Wakade, Bhagyashree Bhame, Deanna A. Hagge, Indra Napit, Mahesh Shah, Neeta Maximus, Joydeepa Darlong, M. Yulianto Listiawan, Shimelis N Doni, Peter Nicholls, Bernd Genser, Saba M. Lambert, Diana N. J. Lockwood, Stephen L. Walker, Erythema Nodosum Leprosum International STudy Group

**Affiliations:** Department of Clinical Research, Faculty of Infectious & Tropical Diseases, London School of Hygiene & Tropical Medicine, London, UK; Department of Dermatology, ALERT Comprehensive Specialized Hospital, Addis Ababa, Ethiopia; Department of Dermatology and Venereology Faculty of Medicine Universitas Airlangga, Dr. Soetomo Teaching Hospital, Surabaya, Indonesia; The Leprosy Mission Trust India, New Delhi, India; Anandaban Hospital, The Leprosy Mission Nepal, Kathmandu, Nepal; Bombay Leprosy Project, Mumbai, India; Zero Leprosy Pathfinder Consulting, Louisiana, USA; ENLIST statistician; High5Data Ltd, Heidelberg, Germany; Medical Faculty Mannheim, University of Heidelberg, Germany

**Keywords:** Leprosy, *Mycobacterium leprae*, erythema nodosum leprosum, type 2 reaction, corticosteroids, severity scale, ENLIST ENL Severity Scale, health-related quality of life

## Abstract

**Background:** Erythema nodosum leprosum (ENL) is a severe inflammatory complication of lepromatous leprosy characterised by recurrent inflammatory episodes often requiring prolonged immunosuppression. The severity of ENL can be quantified using the validated and reliable ENLIST ENL Severity Scale (EESS). The longitudinal course of ENL and how it is captured using standardised severity measures has not been well described. We prospectively evaluated the changes in ENL severity over time using the EESS in a randomised clinical trial.

**Methods:** We conducted a post-hoc analysis of participants enrolled in the Methotrexate and Prednisolone Study in ENL, an international multicentre randomised controlled trial conducted in Ethiopia, India, Indonesia, and Nepal. Adults with severe ENL (EESS score ≥9) were followed for 60 weeks with repeated EESS assessments. Longitudinal trajectories were analysed using mixed-effects regression models. Item-level analyses characterised the clinical phenotype captured by the scale. Associations between EESS score, prednisolone exposure, and dermatology-specific health-related quality of life measured using the Dermatology Life Quality Index (DLQI) were examined.

**Findings:** A total of 135 participants contributed 1,958 EESS assessments. Mean EESS declined rapidly during the first four weeks of treatment (−2.10 points/week; 95% CI −2.36 to −1.84; p<0.001), increased modestly during reduction in corticosteroid dose (weeks 4–20), and gradually declined thereafter. Severe ENL (EESS score ≥9) occurred in 20.6% of visits and was characterised primarily by pain and cutaneous inflammatory manifestations. Participants who required additional prednisolone had persistently higher EESS scores and showed limited improvement compared with those who did not receive additional prednisolone. Longitudinal EESS scores were strongly correlated with the DLQI score (Spearman’s ρ=0.75; p<0.001).

**Conclusion:** The EESS captures clinically meaningful changes in ENL severity, aligns with treatment decisions, and reflects patient-reported severity over time. These findings support the use of the EESS as a robust tool for monitoring ENL severity in both clinical research and routine care.

**Funding:** This work was supported by The Hospital and Homes of St. Giles, grant number ITCRZM25 and Leprosy Research Initiative, Turing Foundation and plan:g under LRI grant number 704.16.71

## INTRODUCTION

Leprosy is a chronic granulomatous disease caused by *Mycobacterium leprae* and *Mycobacterium lepromatosis* (1,2). In 2024, the World Health Organization reported 172,717 new cases globally, with nearly 80% occurring in India, Brazil, and Indonesia (3). Despite effective anti-microbial multidrug therapy (MDT), the clinical course of leprosy is frequently complicated by immune-mediated inflammatory episodes known as leprosy reactions(1). These reactions may occur before, during, or after completion of antimicrobial treatment and are a major cause of morbidity and disability (4,5).

Erythema nodosum leprosum (ENL), also referred to as leprosy Type 2 reaction, is a severe systemic inflammatory complication affecting individuals with lepromatous disease(4). ENL occurs in up to 50% of individuals with lepromatous leprosy and approximately 10% of those with borderline lepromatous leprosy, particularly those with high bacterial indices(6). Clinically, ENL is characterised by recurrent crops of painful erythematous nodules accompanied by peripheral oedema, fever, arthritis, neuritis, bone pain, iritis and orchitis(7). The condition often follows a chronic or recurrent course, contributing substantially to disability, impaired health-related quality of life (HRQoL), and catastrophic household costs (8–11).

Accurate measurement of ENL severity has historically been challenging. Prior to 2016, severity assessment relied largely on subjective clinician judgement or non-validated composite scales (12). The Erythema Nodosum Leprosum International STudy (ENLIST) Group developed and validated the 10-item ENLIST ENL Severity Scale (EESS) in a multicentre study involving 447 participants (13). The EESS generates a score from 0 to 30, with a threshold of ≥9 indicating severe disease (13). Since its validation, the scale has been incorporated into research studies and clinical guidance (14–17); however, published data examining its properties in longitudinal follow-up remain limited(18).

Understanding how the EESS performs over time, how its individual components contribute to disease phenotype, and how changes in score relate to treatment and patient-reported outcomes is critical. A severity instrument may demonstrate reliability and construct validity at a single timepoint yet behave differently with repeated measurements or real-world settings, particularly in chronic inflammatory conditions characterised by fluctuating severity and modified by treatment.

The Methotrexate and Prednisolone Study in ENL (MaPs in ENL) was a multicentre, double-blind, randomised controlled trial conducted in five leprosy referral centres in Ethiopia, India, Indonesia, and Nepal (16). Participants were followed for 60 weeks with repeated EESS assessments alongside detailed data collection on treatment and patient-reported outcome measures including the Dermatology Life Quality Index (DLQI) (16).

In this study, we aimed to characterise changes in ENL severity over time, examine the contribution of individual EESS components, assess responsiveness to treatment, and compare these findings with patient-reported HRQoL using the DLQI in the prospective multicentre MaPs in ENL cohort. Using repeated measures over 60 weeks, we describe how ENL severity changes during treatment and how EESS scores relate to HRQoL.

## METHODS

### Study Design and Participants

We conducted a prespecified secondary longitudinal analysis of the MaPs in ENL trial, a multicentre double-blind randomised controlled trial conducted between 2^nd^ January 2023 and 1^st^ September 2025 at five leprosy referral centres in Ethiopia, India (two sites), Indonesia, and Nepal.

MaPs in ENL enrolled adults with severe ENL, defined as an EESS score of 9 or greater. Participants were followed prospectively for 60 weeks after randomisation with repeated clinical assessments including EESS scores.

The present analysis examined the longitudinal behaviour of ENL severity, focusing on trajectories of EESS scores, the contribution of individual scale components, and the relationship between inflammatory severity and corticosteroid exposure. Treatment allocation in the parent trial was not considered in this analysis.

Ethical approval for the MaPs in ENL trial was obtained from the London School of Hygiene & Tropical Medicine Research Ethics Committee and from ethics committees at all participating study sites, and all participants provided written informed consent before enrolment.

### Trial Procedures

All participants received a standardised oral prednisolone regimen beginning at 40mg daily, tapered to zero over 20 weeks according to a predefined schedule (16).

ENL flares during follow-up were managed according to the trial protocol (16). A flare was defined as clinical worsening of ENL requiring additional prednisolone, based on ENL-related symptoms or signs meeting at least one of the following criteria: increase in EESS score to ≥ 9, or an increase in EESS score by ≥ 5 points from the previous assessment. When a flare occurred, additional prednisolone was prescribed as specified in the protocol(16).

### ENL Severity Assessment

ENL severity was measured using EESS (Table 1), a validated 10-item clinician-administered instrument (13). The scale captures inflammatory and systemic manifestations of ENL defined in the User Guide(13), including pain assessed by visual analogue scale (VAS pain), fever, cutaneous lesion parameters (number, inflammation, and extent of ENL lesions), peripheral oedema, bone pain, joint inflammation attributable to ENL, lymphadenopathy, and nerve tenderness. Each item is scored according to predefined criteria with a score of zero, one, two or three, yielding a total score ranging from 0 to 30. Higher scores indicate greater ENL severity. Severe ENL is defined as EESS score ≥9, as established in the original validation study (13).

**Table 1:**
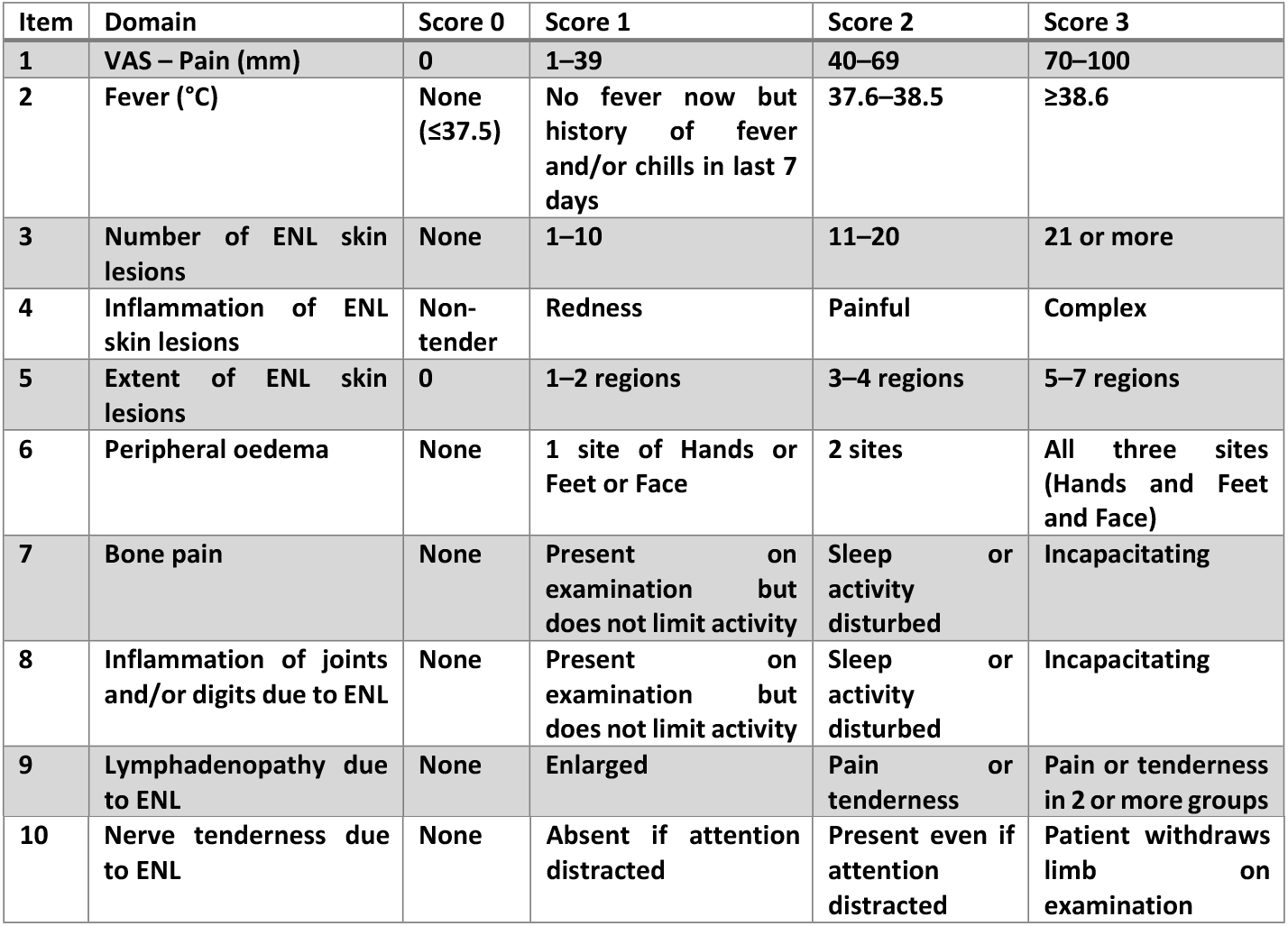
ENLIST ENL Severity Scale scoring system.

### Health Related Quality of Life Assessment

HRQoL was assessed using the DLQI, a validated 10-item patient-reported outcome measure that captures the impact of skin disease on symptoms and feelings, daily activities, leisure, work or school, personal relationships, and treatment during the preceding week. In MaPs in ENL, DLQI was recorded at enrolment and at weeks 24, 48 and 60; it was therefore used to examine the relationship between EESS and HRQoL at matched study timepoints (19).

### Statistical Analysis

The analysed population included participants with at least one valid EESS assessment between randomisation and 60 weeks of follow-up. Longitudinal trajectories of ENL severity were examined using all available repeated EESS measurements. Mean EESS scores were calculated at each study week, and temporal patterns were visualised using locally weighted scatterplot smoothing (LOWESS). Changes in EESS over time were evaluated using mixed-effects linear regression models with random intercepts for participants to account for within-person clustering. Time since randomisation was modelled using piecewise segments representing the early treatment phase (weeks 0–4), corticosteroid taper (weeks 4–20), and post-taper follow-up (weeks 20–60).

To characterise the clinical phenotype captured by the scale, individual EESS components were analysed at both visit and participant level. The proportion of visits in which each component was present was calculated across follow-up, and the proportion of participants experiencing each component at least once was also summarised.

To examine the relationship between ENL severity and treatment requirements during the protocol-defined 20-week prednisolone taper, participants were classified according to whether additional prednisolone was required. Linearity of EESS trajectories over time was assessed using LOWESS plots. As non-linearity was observed, piecewise linear mixed-effects models were fitted with cut-points based on inflection points in the LOWESS curves. Model fit was compared using Akaike’s and Bayesian information criteria, and the piecewise model was retained. Differences between groups were evaluated using mixed-effects models including time, additional prednisolone requirement, and their interaction.

Associations between EESS and DLQI scores were assessed using Spearman’s correlation and mixed-effects regression models. Severe ENL was defined as EESS ≥9. All tests were two-sided and results are presented with 95% confidence intervals. All analyses were conducted using Stata version 18 (StataCorp, College Station, TX, USA).

## RESULTS

Of 137 randomised participants, 135 (98.5%) contributed at least one post-randomisation EESS assessment between 0 and 60 weeks and were included in the longitudinal analysis. These participants contributed 1,958 visits (mean 14.5 visits per participant; range 1–24). Participants had a median age of 33 years (IQR 25–41); 74% were male and 72% had recurrent or chronic ENL at enrolment. Table 2 summarises participants characteristics at enrolment.

**Table 2:**
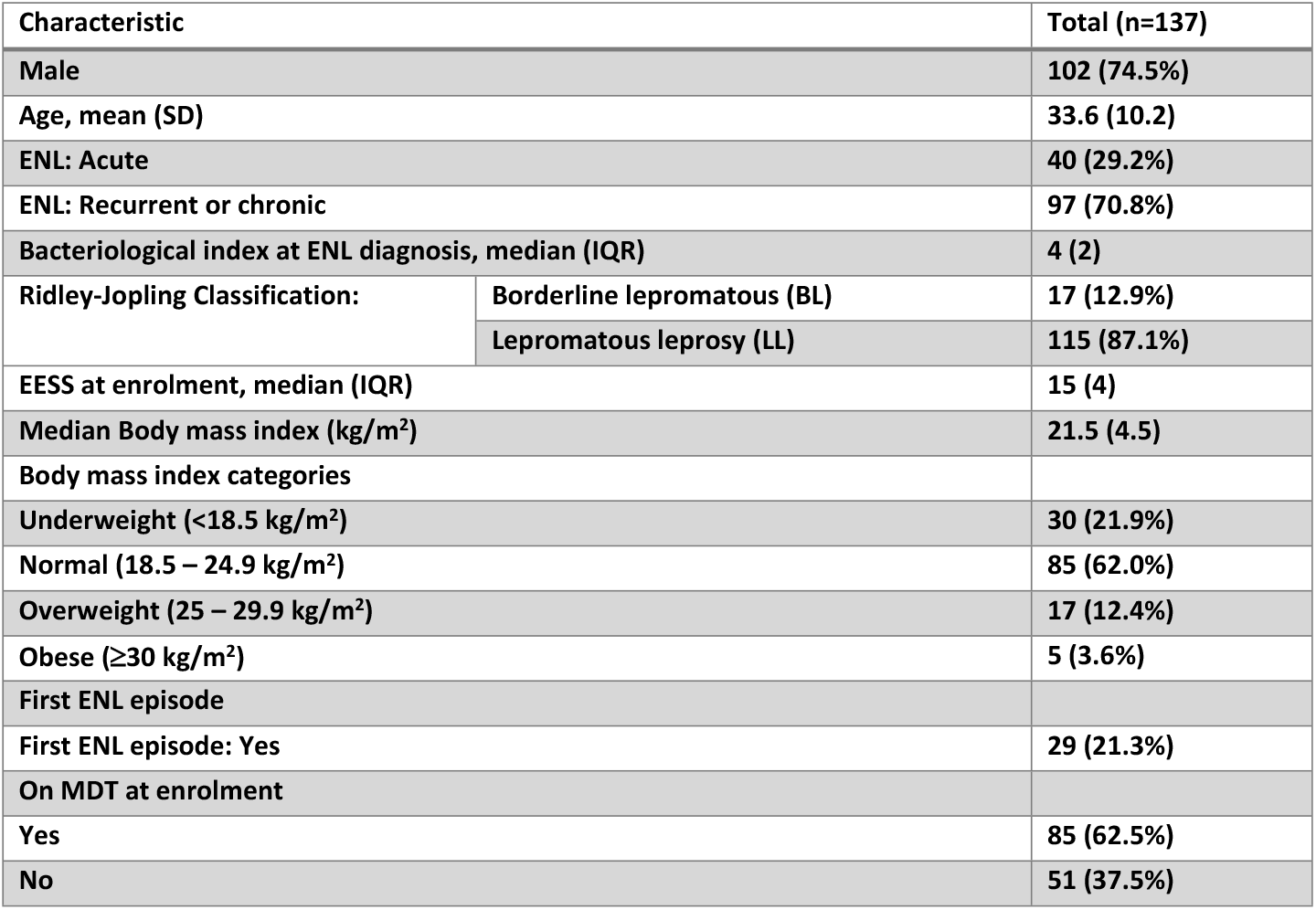
Characteristics of MaPs in ENL cohort.

At enrolment (first recorded assessment), all participants met the protocol-defined severity threshold (EESS ≥9). The mean EESS at enrolment was 14.9 (SD 3.0), with a median of 15 (IQR 13–17).

### Longitudinal analysis of ENL severity

Mean EESS declined sharply during the first four weeks following randomisation, consistent with early corticosteroid response (Figure 1). In piecewise mixed-effects modelling, EESS decreased by 2.10 points per week during weeks 0–4 (95% CI −2.36 to −1.84; p<0.001).

**Figure 1:**
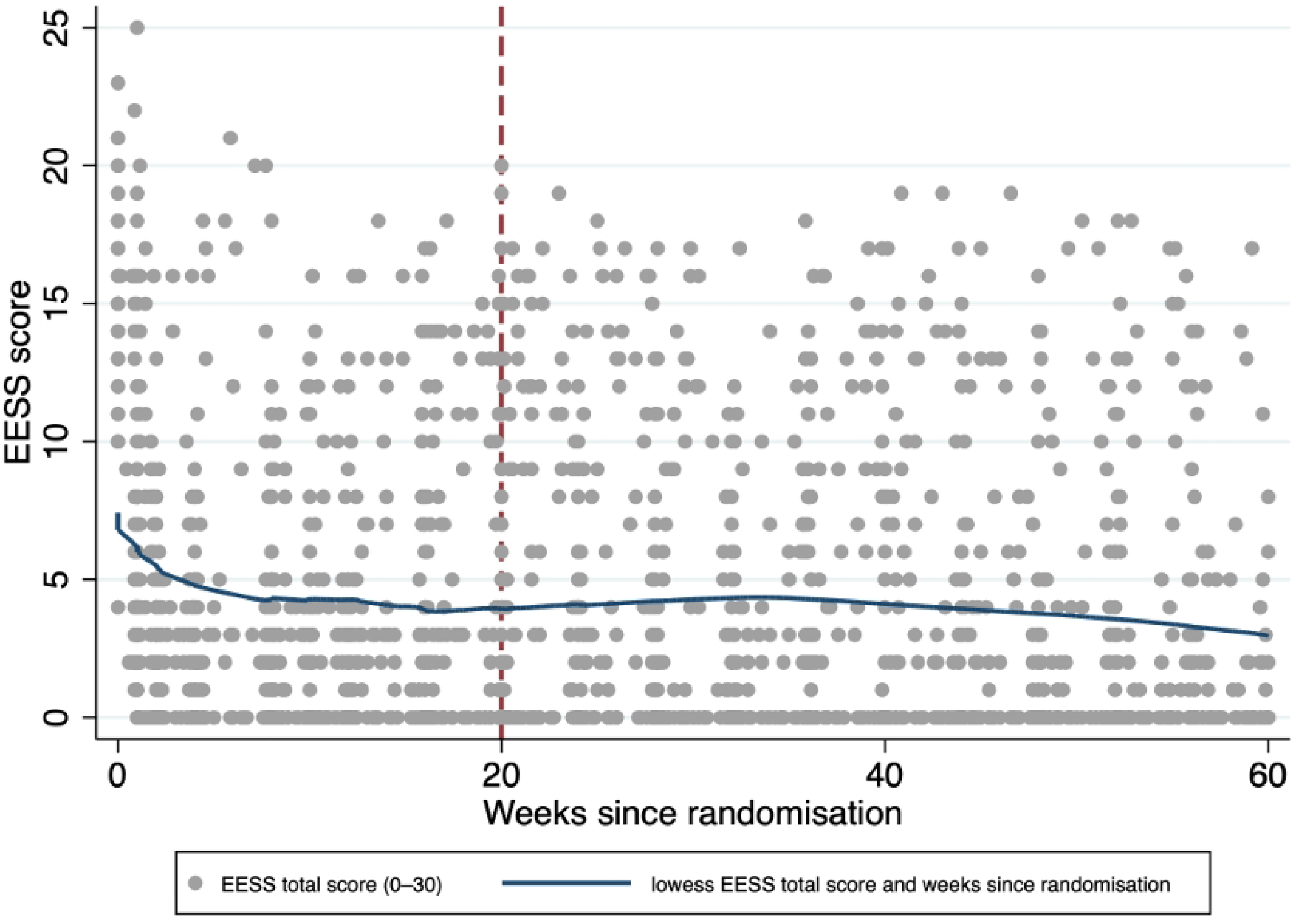
Mean ENLIST ENL Severity Scale score trajectories over 60 weeks. The vertical red dashed line indicates the end of the prednisolone component of the trial intervention.

Between weeks 4 and 20, mean EESS increased modestly (+0.25 points per week; 95% CI 0.20 to 0.30; p<0.001). From weeks 20 to 60, EESS declined gradually (−0.047 points per week; 95% CI −0.070 to −0.025; p<0.001).

Across all visits, EESS scores ranged from 0 to 25 (possible range 0–30). Thirty-eight percent of visits recorded a score of 0, reflecting periods of quiescent ENL. The distribution was right-skewed (skewness 1.10), consistent with fluctuating inflammatory activity over time.

### Clinical phenotype captured by EESS

#### EESS assessment frequencies

Of the 1,958 EESS assessments, VAS pain was present (score ≥1) in 51.3% of EESS assessments. Cutaneous components scoring one or more were present in approximately one-third of assessments: number of ENL skin lesions (36.8%), extent of ENL skin lesions involving one or more regions (35.9%), and inflammation of ENL skin lesions (32.1%). Inflamed joints were recorded in 36.5% of visits, fever in 28.8%, and bone pain in 28.2%. Peripheral oedema occurred in 18.3% of visits. Lymphadenopathy (6.0%) and nerve tenderness (5.4%) were uncommon. The distribution of severity for the principal EESS components over time is shown in Figure 2.

**Figure 2:**
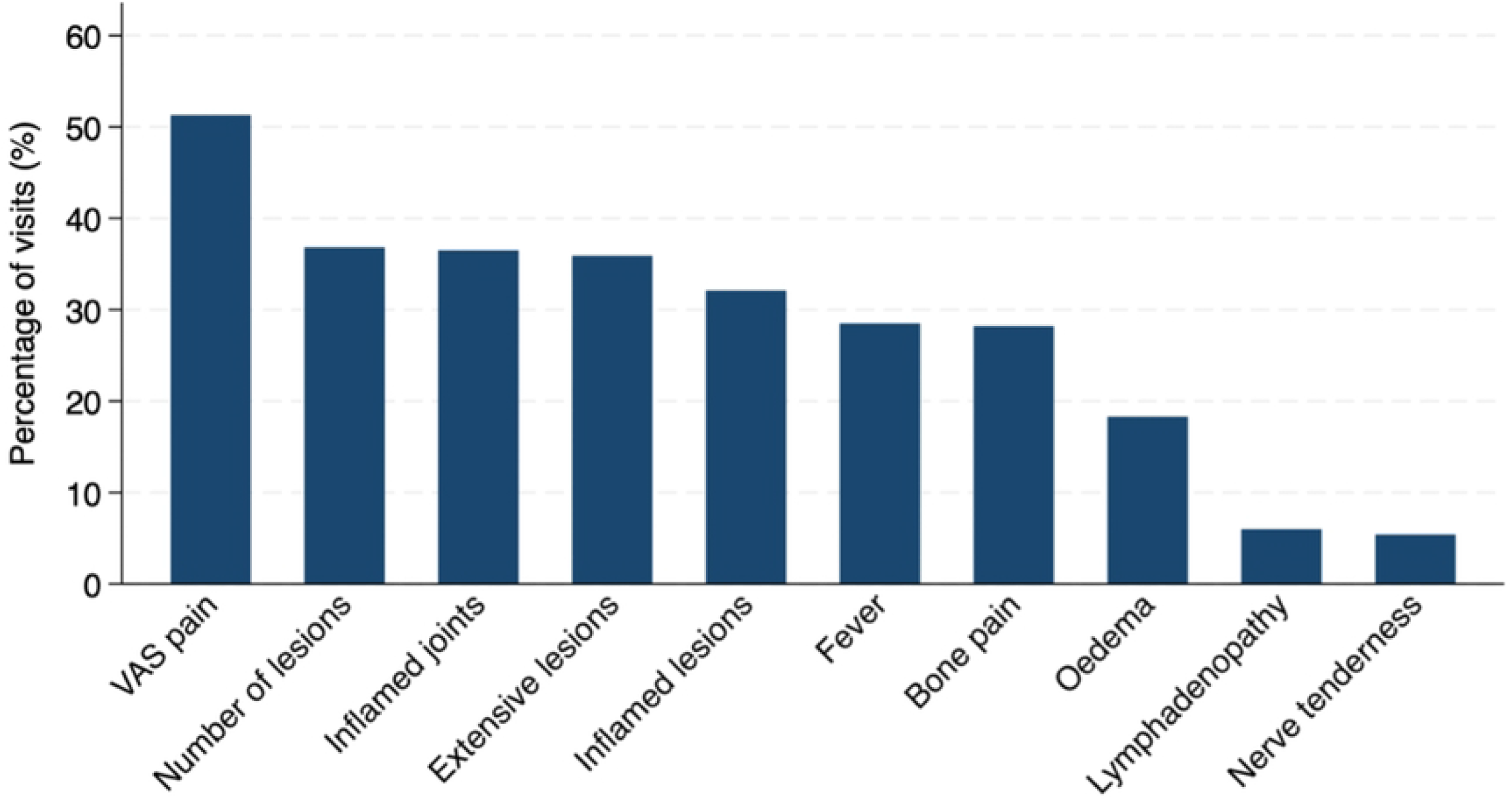
EESS assessment frequency of ENLIST ENL Severity Scale components (0-60 weeks).

The distribution of severity for individual EESS components across all study EESS assessment is shown in Figure 3.

**Figure 3:**
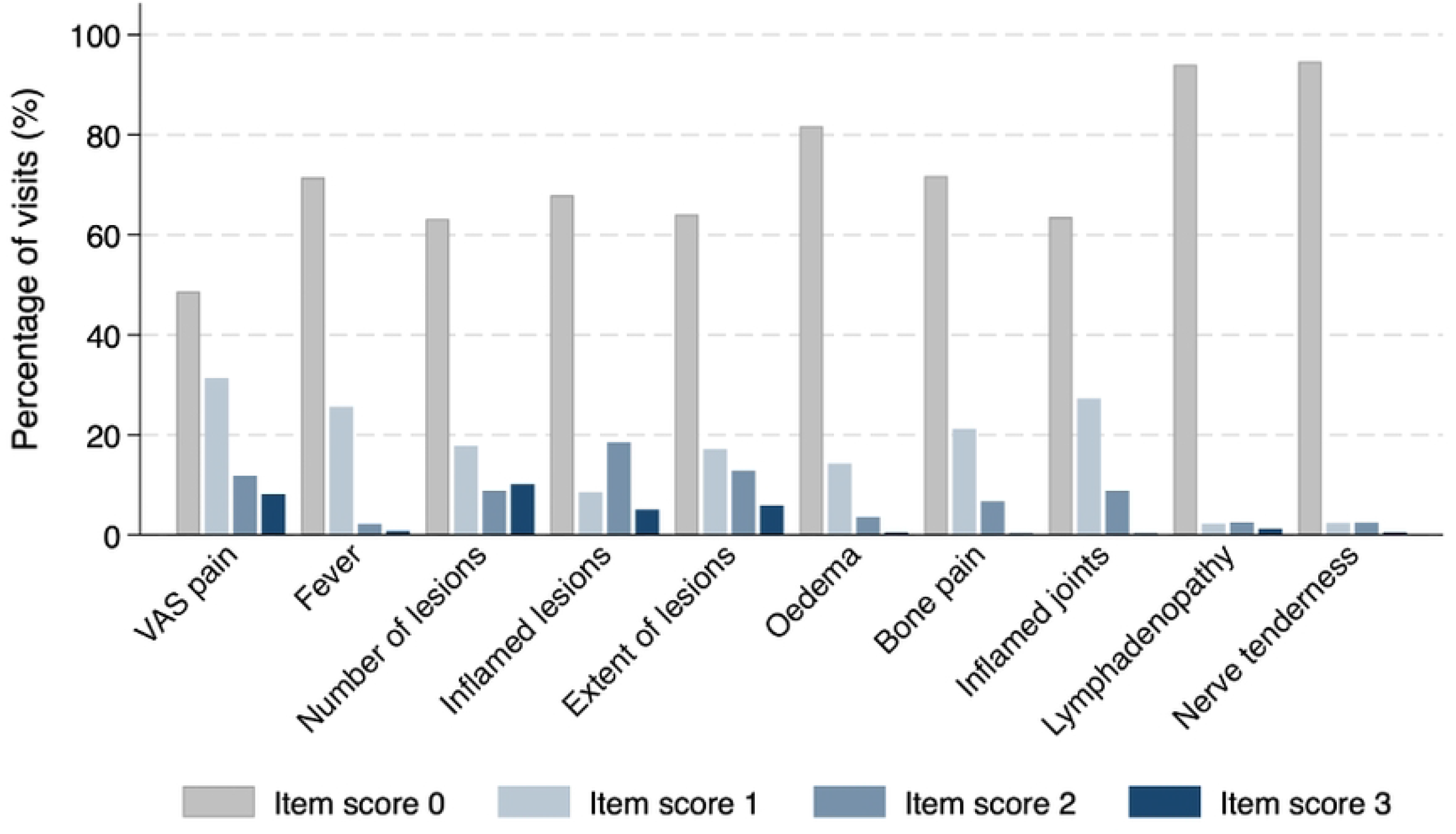
ENLIST ENL Severity Scale components severity frequency

To examine how individual EESS items contributed to overall severity during follow-up, the distribution of scores (0–3) over time was evaluated for five items: pain, number of ENL skin lesions, inflammation of ENL skin lesions, extent of ENL skin lesions, and inflamed joints (Figure 4).

**Figure 4:**
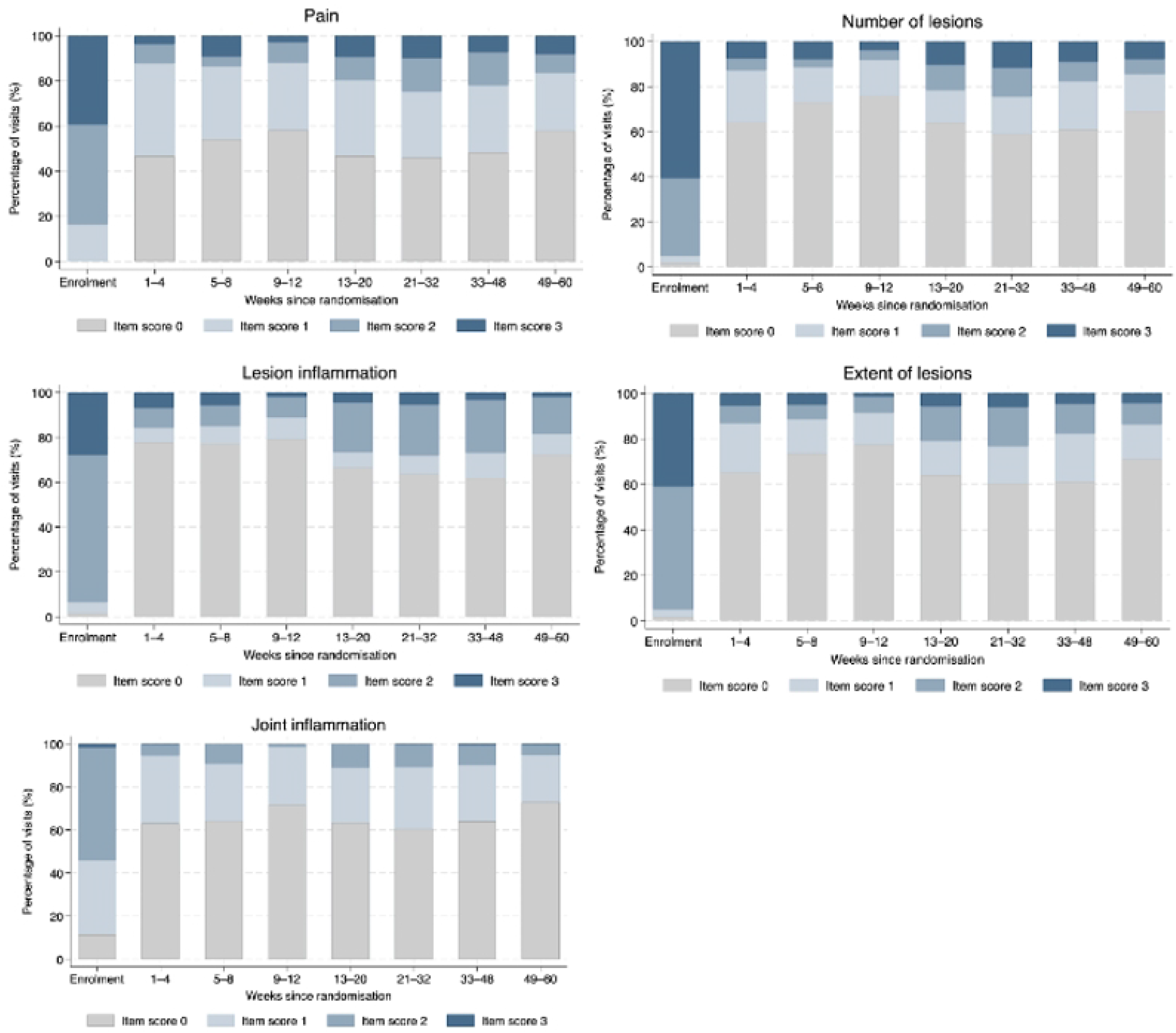
Temporal distribution of key ENLIST ENL Severity Scale components. Stacked bars show the proportion of study visits with scores 0, 1, 2 or 3 for five major scale components across follow-up intervals from randomisation to 60 weeks.

At enrolment, higher scores were common across all items (n=135 assessments). For pain, score 2 and score 3 were observed in 42/135 (31.1%) and 32/135 (23.7%) assessments, respectively. For number of ENL skin lesions, score 2 was present in 35/135 (25.9%) and score 3 in 57/135 (42.2%). For inflammation of ENL skin lesions, score 2 and score 3 were observed in 58/135 (43.0%) and 32/135 (23.7%) assessments, respectively, and for extent of ENL skin lesions in 54/135 (40.0%) and 37/135 (27.4%). For inflamed joints, score 2 was observed in 43/135 (31.9%) assessments.

During weeks 1–4, lower scores were more frequent (n=266 visits). For pain, score 0 and score 1 were observed in 144/266 (54.1%) and 101/266 (38.0%) visits, respectively. For number of ENL skin lesions, score 0 was present in 204/266 (76.7%) and score 1 in 51/266 (19.2%). For inflammation of ENL skin lesions and extent of ENL skin lesions, score 0 was observed in 231/266 (86.8%) and 207/266 (77.8%) visits, respectively. Inflamed joints were predominantly score 0 in 189/266 (71.1%) or score 1 in 70/266 (26.3%).

Between weeks 21–32, higher scores were again observed (n=334 visits). For pain, score 2 and score 3 were present in 50/334 (15.0%) and 33/334 (9.9%) visits. For number of ENL skin lesions, score 2 and score 3 were observed in 43/334 (12.9%) and 39/334 (11.7%), and for inflammation of ENL skin lesions in 76/334 (22.8%) and 18/334 (5.4%). For extent of ENL skin lesions, score 2 and score 3 were observed in 58/334 (17.4%) and 20/334 (6.0%) visits. For inflamed joints, score 2 was observed in 35/334 (10.5%) visits, while score 3 remained ≤0.3% across timepoints.

Overall, these findings indicate that longitudinal changes in ENL severity were primarily driven by pain and cutaneous components, whilst joint involvement contributed less consistently over time.

### Participant-level distribution

At the participant level, defined as experiencing each component at least once during follow-up, eight of the 10 EESS components were recorded in a high proportion of participants. VAS pain was reported by 97.0% of participants. Cutaneous manifestations were also common, including presence of ENL skin lesions (97.8%), involvement of one or more body regions (97.8%), and inflammation of ENL skin lesions (94.1%). Inflamed joints were experienced by 94.8% of participants, fever (current or within the preceding seven days) by 91.9%, bone pain by 89.6%, and peripheral oedema by 78.5%. In contrast, lymphadenopathy (35.6%) and nerve tenderness (31.1%) were less frequently recorded.

These findings indicate that ENL activity captured by the EESS in this cohort was predominantly characterised by inflammatory cutaneous and pain-related features, with nerve tenderness contributing less frequently to overall disease burden but still affecting more than 30% of participants.

### Severe ENL (EESS ≥9)

Across 1,958 post-randomisation assessments, 404 assessments (20.6%) met the threshold for severe ENL (EESS ≥9). VAS pain was present in 99.5% of severe assessments scoring 9 or more, increased lesion number in 99.5%, extensive lesions in 99.3%, and inflamed lesions in 97.5%. Fever, inflammation of joints, and bone pain were substantially more frequent during severe compared with non-severe visits (90.1% vs 12.6%, 86.6% vs 23.4%, and 83.7% vs 13.8%, respectively).

In contrast, lymphadenopathy (17.6%) and nerve tenderness (11.1%) were less common even during severe episodes. These findings indicate that severe ENL was primarily characterised by cutaneous components, fever and pain.

### Relationship between ENL severity and Health-related Quality of Life

At enrolment, the cross-sectional association between EESS and DLQI was weak and not statistically significant (Spearman’s ρ=0.12; p=0.17).

In contrast, longitudinal analyses at matched timepoints (24, 48, and 60 weeks; n=179 observations from 94 participants) demonstrated a strong correlation between ENL severity and the DLQI (Spearman’s ρ=0.75; p<0.001).

In mixed-effects linear regression models accounting for within-participant clustering, each one-point increase in EESS was associated with a 0.92-point increase in DLQI (95% CI 0.80– 1.04; p<0.001). A clinically meaningful five-point increase in EESS corresponded to an approximate 4–5-point increase in DLQI, which is similar to or exceeds the commonly accepted minimal important difference for the DLQI of 4 points.

Severe ENL (EESS ≥9) was associated with a 12-point higher DLQI score compared with non-severe visits (p<0.001). After adjustment for EESS in multivariable models, clinician-defined flare status was not independently associated with DLQI (p=0.15), indicating that HRQoL impairment was primarily explained by measured ENL severity rather than flare classification alone.

### ENL Type and Longitudinal Severity Trajectories

At enrolment (first recorded assessment), the mean EESS score was slightly higher amongst participants with acute ENL (15.6, SD 2.8) compared with those with recurrent or chronic ENL (14.6, SD 3.0), although the difference was small (<1 point on a 30-point scale).

When stratified by ENL type, both acute and recurrent or chronic ENL demonstrated a similar marked decline in EESS during the first four weeks following randomisation, consistent with response to treatment. During the tapering phase (weeks 4–20), inflammatory activity persisted in both groups (Figure 5).

**Figure 5:**
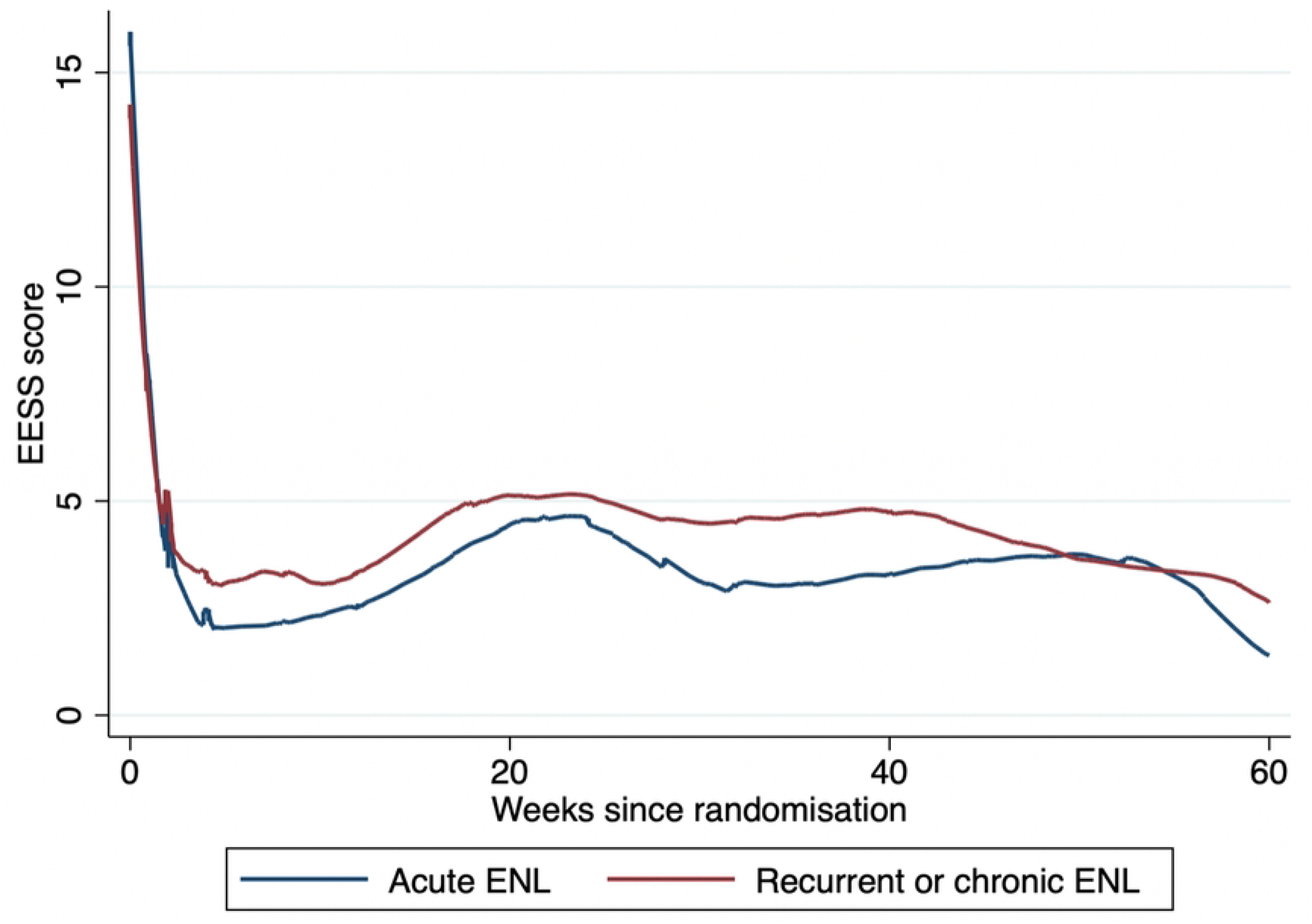
Lowess-smoothed trajectories of ENLIST ENL Severity Scale scores by erythema nodosum type

In mixed-effects models including ENL type as a covariate, recurrent or chronic ENL was associated with a modest increase in mean EESS score during follow-up (β 0.49, 95% CI −0.10 to 1.07; p=0.10), although this difference did not reach statistical significance. There was no strong evidence that early treatment response differed by ENL type.

### EESS scores and treatment

During the 20-week prednisolone taper, EESS score trajectories differed according to whether participants required additional prednisolone. LOWESS plots demonstrated non-linear trajectories, with a rapid early decline followed by a plateau and subsequent increase. In piecewise mixed-effects models, EESS decreased markedly during the first 4 weeks (−2.26 points/week, p<0.001), with a reduced early response among those requiring additional prednisolone (interaction +2.61, p=0.006). From week 4 onwards, EESS scores increased modestly over time (+0.17 points/week, p<0.001), with no difference between groups (Figure 6).

**Figure 6:**
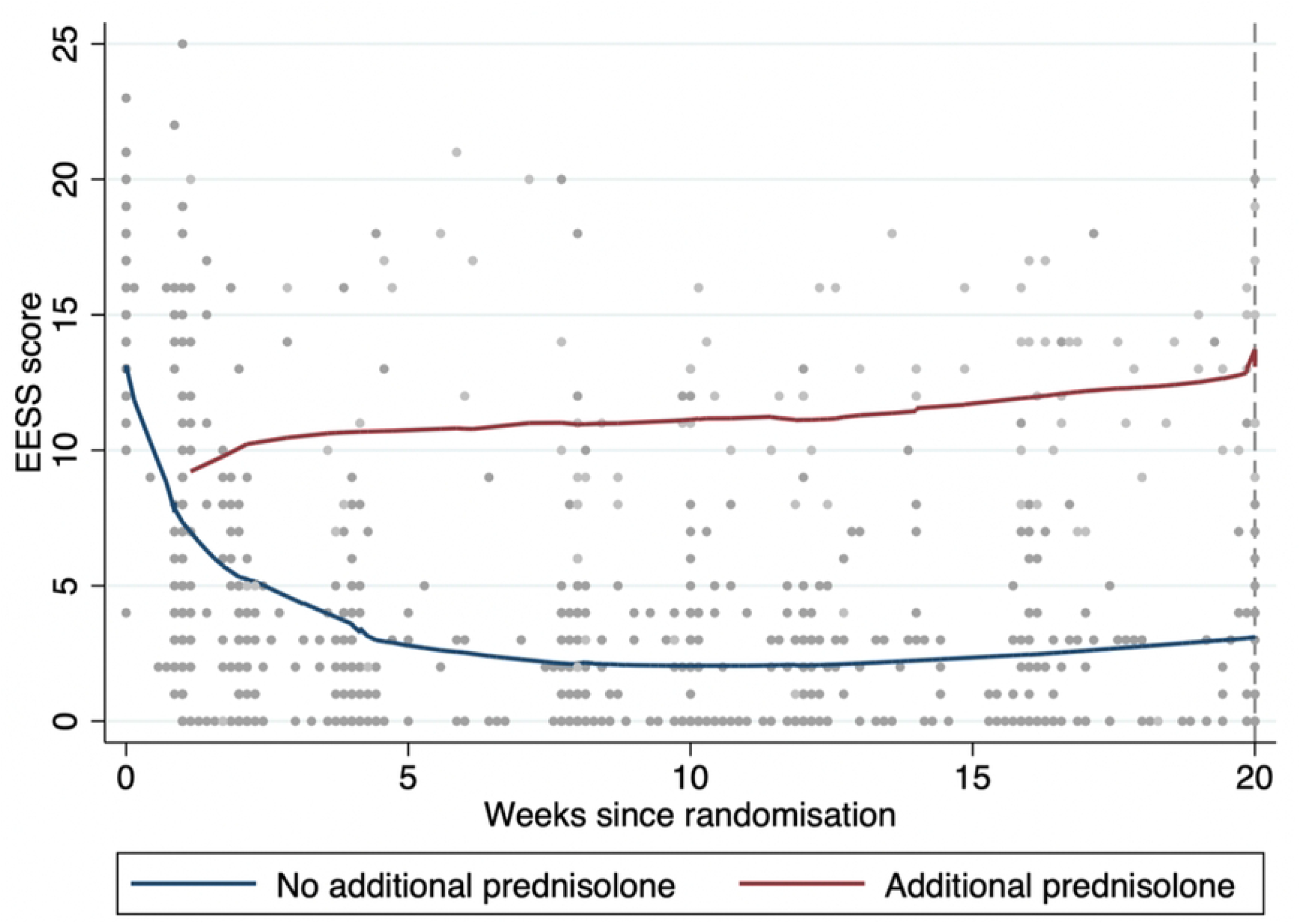
LOWESS-smoothed trajectories of EESS during the first 20 weeks following randomisation. Participants who did not require additional prednisolone demonstrated rapid improvement in ENL severity, whereas those requiring treatment escalation exhibited persistently higher EESS scores and minimal improvement over time.

## DISCUSSION

This study provides a detailed longitudinal evaluation of the EESS in participants enrolled in a large multicentre clinical trial of ENL treatment. Several important observations emerge regarding the behaviour of the scale over time, the contribution of individual clinical domains, and its relationship with treatment response and patient-reported outcomes.

First, the EESS demonstrated a clear temporal pattern during follow-up. Mean scores declined sharply during the first four weeks following the start of treatment, consistent with the expected response to high-dose corticosteroid therapy. After this initial improvement, EESS scores stabilised during the reduction of prednisolone and subsequent follow-up. This pattern reflects the clinical course commonly observed in ENL, in which early suppression of inflammation is achieved with corticosteroids, but disease activity often persists or recurs during dose reduction. During the first 20 weeks after enrolment, corresponding to the planned taper of oral prednisolone, modelled EESS trajectories showed a progressive decline in participants who did not require additional prednisolone. In contrast, participants who required additional prednisolone had persistently higher EESS scores and a reduced early response with subsequent worsening over time, indicating ongoing ENL activity despite treatment. These findings suggest that a smaller early reduction in EESS may be associated with subsequent ENL flares requiring additional prednisolone.

Analysis of individual components of the scale showed that cutaneous components were the most frequent contributors to EESS scores over time. In particular, the number of ENL skin lesions, extent of ENL skin lesions, and inflammation of ENL skin lesions accounted for a substantial proportion of higher scores during follow-up. In contrast, inflammation of joints, lymphadenopathy, and nerve tenderness were less frequently represented by higher scores. These findings indicate that ENL severity, as measured by the EESS, was primarily driven by cutaneous components and pain over time.

EESS scores correlated with patient-reported quality of life. ENL episodes with EESS scores ≥9 were associated with substantially higher DLQI scores, indicating marked impairment in daily functioning.

Acute and recurrent or chronic ENL demonstrated broadly similar trajectories over time. Although recurrent or chronic ENL was associated with slightly higher EESS scores during follow-up, the difference was modest and did not reach statistical significance.

Our findings are consistent with previous studies evaluating the EESS in smaller cohorts. In a retrospective study of eight patients with ENL in Hawaii (74 observations over three years), longitudinal assessment showed that EESS scores were largely influenced by the three cutaneous items (18). The present study confirms these observations in a substantially larger, more diverse population with prospectively collected data.

The EESS has also been applied in biomarker studies of ENL. In a cross-sectional study, serum interleukin-6 (IL-6) levels were strongly positively correlated with EESS scores (r=0.81, p<0.05), with higher IL-6 levels observed in individuals with more severe EESS scores, suggesting that EESS scores correlate with inflammation markers (20). The use of a standardised severity instrument is essential in clinical studies and may have a role in the validation of biomarkers and improve the design of pathogenesis studies.

The analysis of the EESS data from the MaPs in ENL supports the use of the EESS as a robust and clinically meaningful measure of ENL severity. The scale captures key domains of inflammatory disease activity, reflects patient-reported outcome measures of HRQoL, and is sensitive to differences in treatment response during corticosteroid therapy. These characteristics reinforce its value as a standardised outcome measure for clinical trials and other studies of ENL and supports its wider incorporation into routine clinical practice to improve the assessment and monitoring of disease severity and treatment decisions.

### Strengths and limitations

This study has important strengths. It draws on prospectively collected data from a multicentre international cohort from four leprosy endemic countries, with frequent and standardised severity assessments over 60 weeks. The longitudinal design and large number of assessments enhance precision in trajectory estimation. Additionally, the analysis reflects real-world disease response using standardised corticosteroid dosing.

However, several limitations warrant consideration. All participants had severe ENL at enrolment, limiting generalisability to milder disease. The study population was derived from a clinical trial setting, which may not fully reflect routine care. Furthermore, whilst EESS captures inflammatory manifestations comprehensively, it does not capture permanent disability or cumulative nerve impairment.

## CONCLUSION

In this multicentre longitudinal cohort of adults with severe ENL, the EESS demonstrated responsiveness to treatment and captured the fluctuating inflammatory course of ENL during treatment with corticosteroids. Disease severity was driven by cutaneous components and pain, particularly the number of ENL skin lesions, extent of ENL skin lesions, and inflammation of ENL skin lesions, while lymphadenopathy and nerve tenderness were less frequently represented by higher scores.

Our findings confirm the EESS is a robust tool for longitudinal monitoring of ENL severity and as a standardised outcome measure for both research and clinical practice.

### SUPPORT

This work was supported by The Hospital and Homes of St. Giles, grant number ITCRZM25 and Leprosy Research Initiative, Turing Foundation and plan:g under LRI grant number 704.16.71

## Data Availability

The de-identified individual participant dataset underlying the results reported in this article, together with the data dictionary and statistical analysis code, will be deposited in the LSHTM Data Compass repository. The dataset will be made publicly available following a one-year embargo after publication of the article to allow completion of planned secondary analyses by the study investigators.

## ACKNOWLEDGEMENTS

We would like to express our gratitude to the people with lived experience of leprosy who participated in, or considered participating in, this study. Their time and commitment made this research possible.

We thank the clinical and research staff at all participating centres for their dedication to recruitment, follow-up, and patient care.

We acknowledge the contribution of the ENLIST Group, including Marivic Balagon, C. Ruth Butlin, Milton O. Moraes (in memoriam), Jose da Costa Nery, and Anna Sales, for their continued commitment to advancing research in ENL and improving outcomes for affected communities.

## AUTHOR CONTRIBUTIONS

Conceptualisation: Barbara de Barros, Diana N. J. Lockwood, Stephen L. Walker and the Erythema Nodosum Leprosum International Study Group (ENLIST)

Methodology: Barbara de Barros, Bernd Genser, Peter Nicholls, Diana N. J. Lockwood and Stephen L. Walker

Statistical Analysis: Barbara de Barros, Stephen L. Walker and Bernd Genser

Investigation: Farha Sultana, Anju Wakade, Bhagyashree Bhame, Bishwanath Acharya, Abdulnaser Hamza, Alemtsehay Getachew, Medhi Denisa Alinda, M. Yulianto Listiawan, Shimelis N Doni, Deana A. Hagge, Indra Napit, Mahesh Shah, Vivek V. Pai, Neeta Maximus, Joydeepa Darlong

Data Curation: Barbara de Barros, Bernd Genser, Peter Nicholls

Project Administration: Barbara de Barros

Supervision: Diana N. J. Lockwood and Stephen L. Walker

Writing – Original Draft Preparation: Barbara de Barros

Writing – Review & Editing: All authors

Funding Acquisition: Diana N. J. Lockwood, Stephen L. Walker

